# Evaluation of the Irritation and Sensitization Potential of Medical-grade Norway Spruce (*Picea abies*) Resin Salve: Single-Blind Modified Draize Repeat Insult Patch Test in Healthy Volunteers

**DOI:** 10.1101/2025.02.20.25320648

**Authors:** Kamilla Yamileva, Evgen Multia

## Abstract

**Background:** Abilar® is a wound salve containing 10 % medical-grade Norway spruce (*Picea abies*) resin, known for its antimicrobial and wound-healing properties. However, isolated reports of allergic contact dermatitis have raised concerns regarding its cutaneous safety profile. Given the increasing use of *Picea abies* resin-based products, it is important to evaluate any potential for skin irritation and sensitization.

**Objectives:** To evaluate the irritation and sensitization potential of medical-grade *Picea abies* resin salve using a modified Draize Human Repeat Insult Patch Test (HRIPT) in a healthy adult cohort, thereby providing toxicological insights relevant to both clinical applications and consumer safety.

**Methods:** A single-blind study (ClinicalTrial.gov, Identifier: NCT06810856) was conducted with 215 healthy volunteers (207 completed the study). Medical-grade *Picea abies* resin salve was applied through cutaneous patches in an induction phase and a challenge phase. Skin reactions were evaluated using modified Draize scoring system, with reactions to both the test material and adhesive being recorded separately.

**Results:** During the induction phase, only 7 of 207 subjects (3.38%) of participants exhibited mild erythema (Grade 1) attributed to the resin salve. Notably, no participant experienced moderate to severe reactions (Grades 2–5). In the subsequent challenge phase, no reactions were observed, and subjects with prior Grade 1 responses reverted to a non-reactive status. These findings indicate that, under the test conditions and repeated exposure regimen, the medical-grade *Picea abies* resin salve demonstrates a low potential for inducing cutaneous irritation or sensitization.

**Conclusions:** The HRIPT findings demonstrate that medical-grade *Picea abies* resin salve has a low irritation and sensitization potential under the conditions tested. Although rare cases of allergic contact dermatitis have been documented in literature in predisposed individuals, the data of this study suggests that the overall risk in the general population is low. These findings are also supported by clinical studies and extensive post-market surveillance of Abilar® in wound care for both acute and chronic wounds.

**Graphical Abstract:** 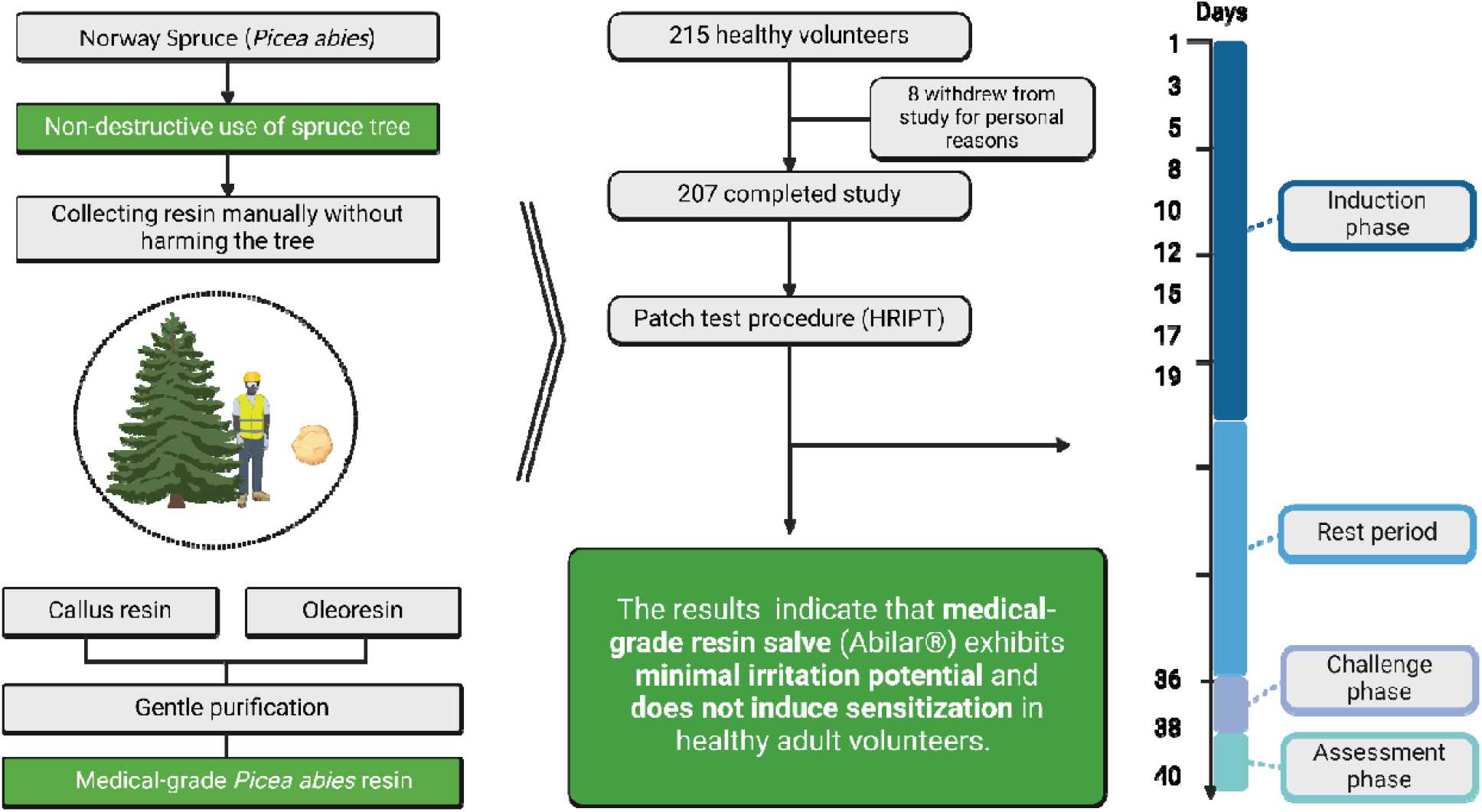

## 1. Introduction

Norway spruce (*Picea abies*) resin is typically found in nature as a mixture of callus and oleoresin and is composed of p-Coumaric acid, resin acids, and lignans (1). The oleoresin part consists mainly of resin acids and forms a protective barrier when the tree is wounded, guarding the spruce tree against dehydration and microbes (2,3). The callus resin forms around the closing wounds of the tree and contains mostly lignans and p-Coumaric acid (1). Together, callus and oleoresin work to protect, seal, and eventually heal the tree’s wound. Beyond its role in tree’s defense, *Picea abies* resin can be also used to treat wounds and various skin conditions in both humans (4) and animals (5). Traditionally, it has been used in Nordic folk medicine to treat a variety of skin diseases, including non-infected and infected wounds, due to its antimicrobial (6) and wound-healing properties (7). Building on this historical foundation, a medical device (Abilar®) was developed in 2008, with clinical testing and years of post-market surveillance confirming its effectiveness and safety in modern medical settings. In this medical device, a purified medical-grade *Picea abies* resin is used. For example, in a prospective, randomized, controlled multicenter trial medical-grade *Picea abies* resin salve demonstrated better efficacy of compared to a sodium carboxy-methylcellulose hydrocolloid polymer in the treatment of severe pressure ulcers, with 92% of patients in the resin group achieving complete healing within six months (8).

Although coniferous resins and rosins are widely used, concerns remain about their allergenic potential (9). Reports in the literature highlight cases of allergies to industrial pine rosin, also known as colophonium (9–12). In Sweden, approximately 5% of patients patch tested for dermatitis symptoms in a special dermatology clinic showed allergy to colophonium (11). However, this figure likely reflects a selection bias, as dermatology clinics typically see patients already predisposed to or exhibiting symptoms of contact dermatitis, which may overestimate the prevalence of colophonium allergy in the general population. In contrast, studies conducted on unselected populations have reported lower prevalence rates. For example, in a study of 793 Danish adults (567 participated) showed a prevalence of colophonium allergy of 0.4% in men (n=279) and 1.0% in women (n=288) (13). In another study of 1236 Norwegian adults, it was found that colophonium allergy rate were 0.9% for men (n=546) and 1.4% for women (n=690), with irritant reaction noticed in 0.2 % (14). These results indicate that the prevalence of allergic reactions to colophonium varies (0.4-5%) but generally remains low.

It is also essential to distinguish between medical-grade *Picea abies* resin from Norway spruce tree, used in medical devices like Abilar®, and colophonium from pine trees (genus *Pinus*). Colophonium is a complex mix of resin acids, with its composition varying by origin and production methods (15,16). Three sources of colophonium can be found: gum rosin (from tapped living pine trees), wood rosin (from old pine stumps), and tall oil rosin (a by-product of pine tree kraft pulping) (17). To produce colophonium, the pine oleoresin must undergo chemical processing to separate the solid resin acids (the colophonium) from the volatile and liquid fractions (e.g., turpentine). The chemical composition of *Picea abies* resin also differs significantly from colophonium. In addition to differing resin acid composition and proportions, colophonium entirely lacks p-*Coumaric acid* and lignans, which are present in medical-grade *Picea abies* resin. Furthermore, medical-grade *Picea abies* resin used for Abilar® is harvested manually from the spruce bark surface without harming the trees and purified without heating. The used process preserves medical-grade *Picea abies* resin’s natural composition and minimize the formation of oxidized resin acids, which are known to be more allergenic.

While the therapeutic benefits of medical-grade *Picea abies* resin are well-documented, studies regarding its allergenic potential are not as well documented apart from clinical studies (4,8,18,19). It is thus important to assess the irritation and sensitization potential of medical-grade *Picea abies* resin salve. While one case report (9) has shown sensitization incidents to Abilar® in polysensitized individuals, large-scale controlled study in healthy population is needed to evaluate the overall safety profile. This study aims to evaluate the irritation and sensitization potential of Abilar® using a modified Draize human repeat insult patch test (HRIPT) on healthy volunteers.

## 2. Patients, Materials and Methods

### 2.1 Study Design

This single-blind, within-subject comparison study was conducted by PCR Corp to assess the irritation and sensitization potential of medical-grade *Picea abies* resin salve (Abilar®). The study followed the modified Draize method of Jordan and King (20), designed for repeated cutaneous patch applications to evaluate claims such as “Dermatologically Tested,” “Clinically Tested,” and “Safe for Skin.” The study was conducted under controlled conditions, adhering to applicable regulatory guidelines for human research.

### 2.2 Participants

A total of 215 healthy volunteers (age range: 18–60 years) were recruited, with 207 completing the study (Figure 1). The protocol aimed at a minimum of 200 subjects to ensure statistical power for assessing the irritation and sensitization potential of medical-grade *Picea abies* resin salve.

**Figure 1:**
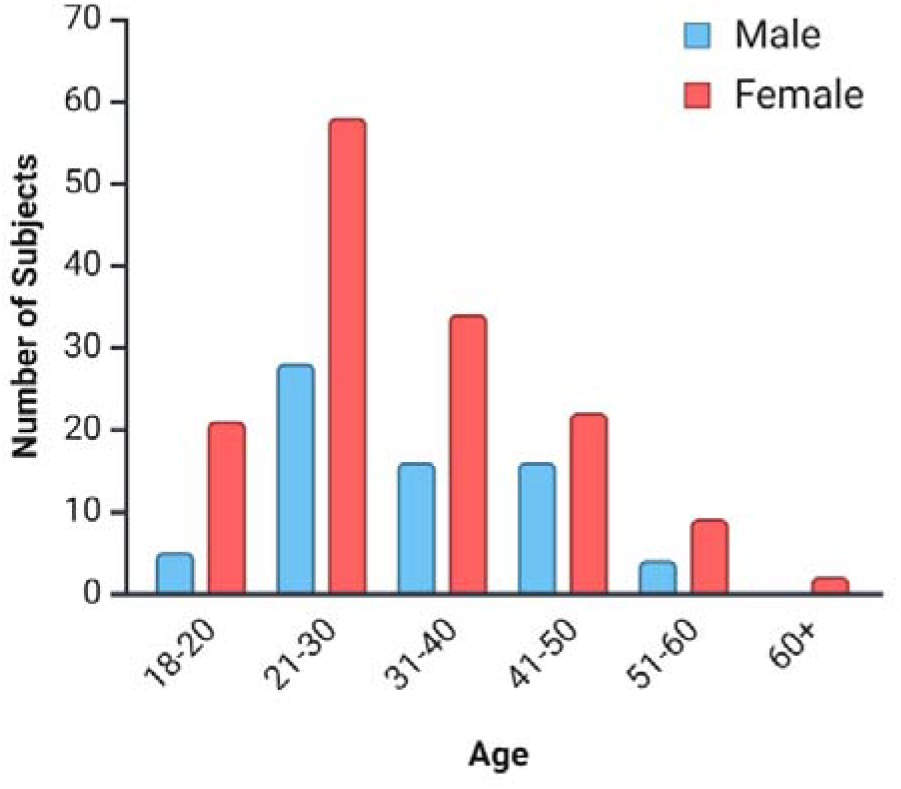
Age and sex distribution of participants.

Both male and female participants were included in the study. Participants were included if they were healthy adults (aged 18 years or older) willing to provide written informed consent. Confounding factors such as prior exposure to rosin-based products or other allergens were carefully controlled through exclusion criteria. For instance, individuals with known sensitivity to rosin or adhesive tape were excluded to ensure that observed reactions could be more confidently attributed to medical-grade *Picea abies* resin (21).

The study was done in a low-risk population. The following exclusion criteria were applied: Pregnancy or lactation. Inadequate or non-existent contraception (women of childbearing potential only). A current skin disease of any type apart from mild facial acne (e.g., eczema, psoriasis).

Heavy alcohol consumption (i.e. more than 21 units per week or 8 units a day for men, more than 14 units per week or 4 units a day for women). Current use or history of repeated use of street drugs. A febrile illness lasting more than 24 hours in the six days prior to first patch application. Significant past medical history of hepatic, renal, cardiac, pulmonary, digestive, hematological, neurological, locomotor or psychiatric disease. History of asthma only if requiring regular medication or hay fever that required prescription treatment in two or more of the previous three years. A history of multiple drug hypersensitivity. Concurrent medication that is likely to affect the response to the test articles or confuse the results of the study. Known sensitivity to the test articles or their constituents including patch materials. Current treatment by a physician for allergy. Participation in a repeat insult patch test (RIPT) or follow-up work within the last month. Sensitization or questionable sensitization in a RIPT. Recent immunization (less than 10 days prior to test patch application). A medical history indicating atopy.

### 2.3 Test article

The test article was a class IIb medical device Abilar® wound salve, containing 10% of medical-grade Norway spruce (*Picea abies*) resin in a salve base. The resin was collected in Finland and extracted in ethanol according to a patented method (22). The resin was purified without heating to preserve its natural composition and minimize the formation of oxidized resin acids, which are known to be more allergenic (23–25). The detailed ingredient names according to International Nomenclature of Cosmetic Ingredients (INCI) are listed in Table 1.

**Table 1.** INCI list of Abilar® ingredients.

| Ingredient | CAS |
| --- | --- |
| Petrolatum | 8009-03-8 |
| Paraffinum Liquidum | 8012-95-1 |
| Norway Spruce (*Picea abies*) Resin |  |
| Ethanol Denat. | 64-17-5 |
| Cera Microcristallina | 63231-60-7 |
| Sorbitan Oleate | 1338-43-8 |
| Cera Alba | 8012-89-3 |
| Hydrogenated Castor Oil | 8001-78-3 |
| Stearic Acid | 57-11-4 |

### 2.4 Patch Test Procedure

Hypoallergenic Scanpor® tape (Norges-plaster A/S) containing no colophony and Finn chambers were used for patch applications. Each patch was 6 cm wide, with Finn chambers fixed to the skin using the tape. Crystal violet dye was applied at the top and bottom of the tape to ensure exact relocation of patches during the repeated application phases.

### 2.5 Induction Phase

Patches containing approximately 0.02 g of medical-grade *Picea abies* resin salve were applied to the upper back of participants on Days 1, 3, 5, 8, 10, 12, 15, 17, and 19. The subjects were instructed to keep the patches in place for 47 hours then to remove and discard them. Patches were applied to the same site each day unless a reaction stronger than mild erythema was present in which case the patch strip was cut and the relevant patches moved outwards. Assessment of patch sites was done immediately before application of the next patch on Days 3, 5, 8, 10, 12, 15, 17, 19 and 22.

### 2.6 Rest Period

Participants were not exposed to patches containing medical-grade *Picea abies* resin salve during Days 22-35. This rest period allowed the immune system time to potentially develop a sensitized response, which can then be detected during the challenge phase. Skin was allowed to recover from any irritation caused by the induction phase.

### 2.7 Challenge Phase

Challenge patches were applied to the upper back (away from the induction site) of each subject on Day 36 to a naïve skin site. Subjects were required to remain at the test center for one hour following application of the patches. The patches were worn for 47 hours and were then removed and discarded by the subject. Challenge assessments were then performed on Days 38 and 40. Assessment of patch sites was at one hour and 49 hours after patch removal.

### 2.8 Assessment

Illumination of the patch sites was done with a 60-watt pearl bulb, approximately 30 cm from the site. The primary outcome was the incidence and severity of skin reactions at the patch sites during both the induction and challenge phases. Reactions were attributed to the test material and to the adhesive tape, both being documented separately. Sensitization, if present, was identified by severe erythema, oedema, papules, and/or vesicles.

Scoring was based on the modified Draize Scale:

- Grade 0 - No visible reaction. This score would include superficial skin responses such as glazing, peeling, cracking.
- Grade 1 - Mild erythematous reaction. Faint pink to definite pink.
  - 1E - Mild erythematous reaction with papules and/or oedema.
- Grade 2 - Moderate erythematous reaction. Definite pink to red erythema (similar to sunburn)
  - 2E - Moderate erythematous reaction with oedema and/or papules.
- Grade 3 - Strong erythematous reaction. Beet red.
  - 3E - Strong erythematous reaction with marked oedema, papules and/or few vesicles.
- Grade 4 - Severe reaction with erythema, oedema, papules and vesicles (may be evidence of weeping).
- Grade 5 - Bullous reaction.

Adhesive-related reactions were recorded separately to distinguish them from reactions caused by medical-grade *Picea abies* resin.

### 2.9 Ethics, registration, and approvals

The study adhered to the principles of the Declaration of Helsinki and its subsequent amendments (26). Ethical approval was granted by the East Anglia Ethics Committee (Princeton Consumer Research Corp, test reference REPRIP2M) for conducting the HRIPT protocol between January 2019 and January 2020. All participants provided written informed consent before participating, and their safety was ensured throughout the study. The study was conducted in line with the ICH Guidelines on Good Clinical Practice (1996) and other recognized international guidelines for human research. The study was retrospectively registered and can be accessed on ClinicalTrials.gov (Identifier: NCT06810856).

## 3 Results

### 3.1 Demographic Data

Out of 215 recruited subjects, 207 completed the study. Eight subjects withdrew from this study for personal reasons. All eight withdrawals showed no reactions prior to their withdrawal. These subjects were not followed up after their withdrawal from study and withdrawn subjects were not replaced.

### 3.2 Reaction Grades Distribution

No serious adverse events or reactions were reported in the study. Only minimal erythema (Grade 1) was observed during the induction phase of the study for some of the participants.

### 3.3 Induction Phase

Adhesive reactions (‘A’) attributed to the adhesive tape were observed in 34 out of 207 participants (16.43%). Only 1 out of 207 participants (0.48%) exhibited a mild erythema reaction that could be attributed solely to resin salve without simultaneous adhesive reactions. 6 out of 207 participants (2.90%) had both adhesive reactions and mild erythema simultaneously. 1 out of 207 participants (0.48%) exhibited a mild erythema reaction week before having an adhesive reaction as well. In total, there were 7 Grade 1 reactions (3.38%). A total of 35 out of 207 participants (16.91%) experienced some form of reaction. 172 out of 207 participants (83.09%) experienced no reactions during the study. The results are collected in Table 2.

**Table 2:** Summary of participant reactions during Induction Phase.

| Reaction Type | Number of Participants | Percentage (%) |
| --- | --- | --- |
| Adhesive Reactions ('A') | 34 | 16.43 |
| Reactions (Grade '1' only) without Simultaneous Adhesive Reactions | 1 | 0.48 |
| Simultaneous Adhesive Reactions 'A' and ' Grade 1' Reactions | 6 | 2.90 |
| Total reactions (Grade 1) | 7 | 3.38 |
| Any Reaction (Adhesive or ' Grade 1') | 35 | 16.91 |
| No Reactions | 172 | 83.09 |
| **Total Participants** | **207** | **100** |

No statistically significant associations were detected between sex or age groups and the likelihood of experiencing a Grade 1 reaction to the medical-grade *Picea abies* resin salve (Table 3). Higher reaction rates observed in males and specific age groups (18-25 and 36-45) were not statistically significant (chi-square test), due to the small number of reactions and limited sample sizes in subgroups. Both males and females exhibited low and comparable reaction rates, indicating that medical-grade *Picea abies* resin salve is generally safe across genders. The lack of significant differences in reaction rates among various age groups suggests that medical-grade *Picea abies* resin salve is suitable for patients of different ages, including the elderly who are more prone to chronic wounds.

**Table 3.**
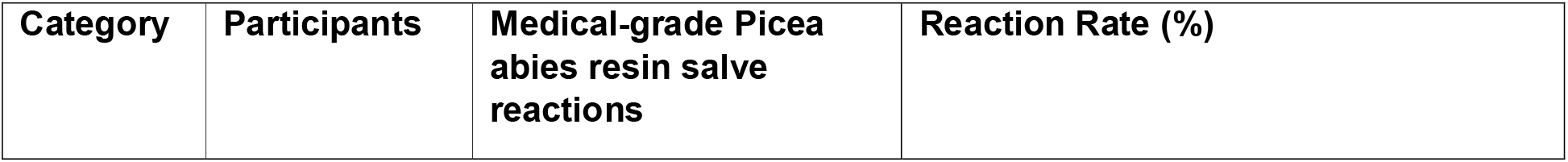
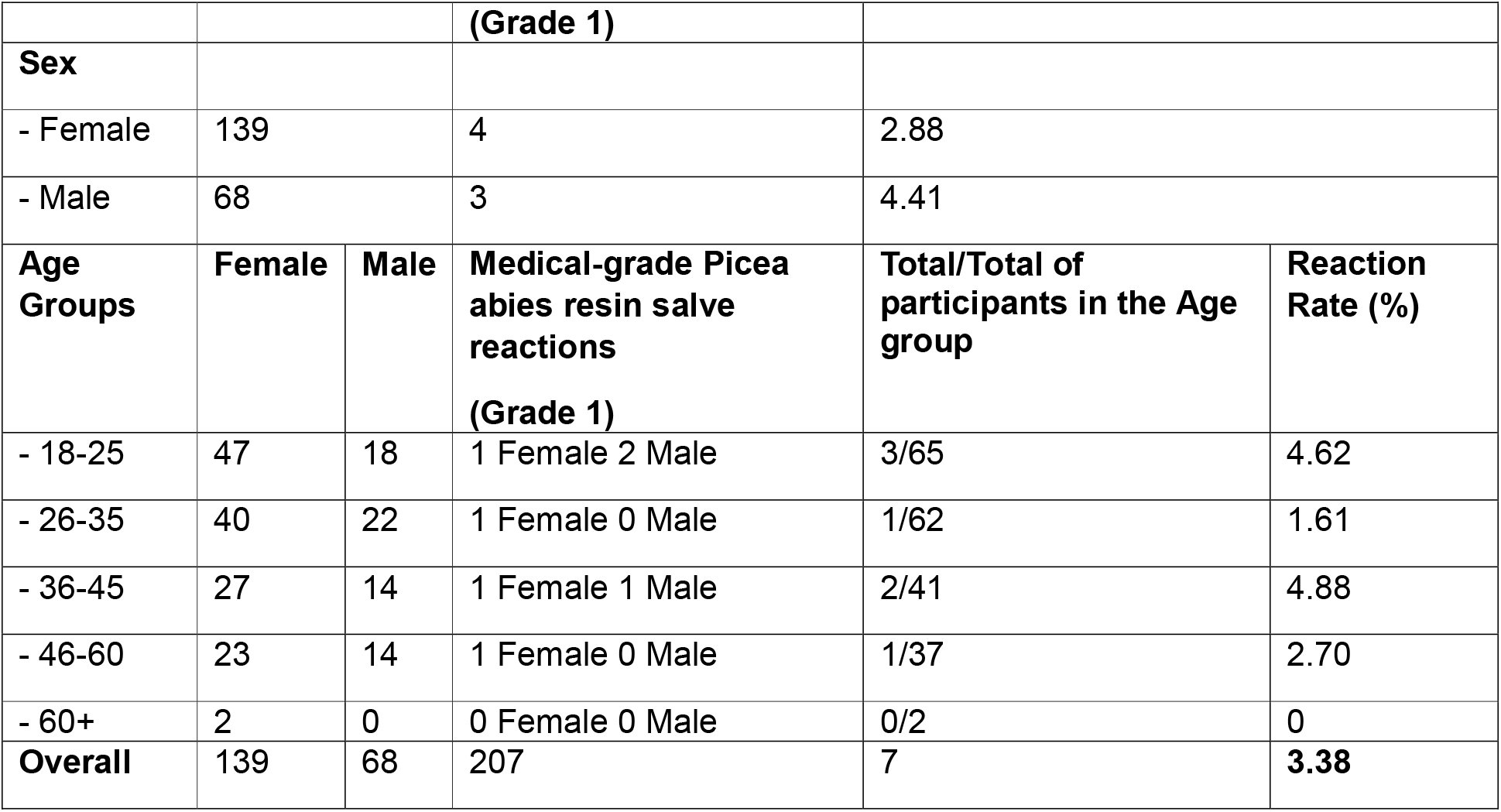
Summary of Sex and Age Group distributions.

| Category | Participants | Medical-grade *Picea abies* resin salve reactions | Reaction Rate (%) |
| --- | --- | --- | --- |

|  |  |  |  |  |  |
| --- | --- | --- | --- | --- | --- |
|  |  |  | (Grade 1) |  |  |
| **Sex** |  |  |  |  |  |
| - Female | 139 |  | 4 | 2.88 |  |
| - Male | 68 |  | 3 | 4.41 |  |
| **Age Groups** | **Female** | **Male** | **Medical-grade Picea abies resin salve reactions (Grade 1)** | **Total/Total of participants in the Age group** | **Reaction Rate (%)** |
| - 18-25 | 47 | 18 | 1 Female 2 Male | 3/65 | 4.62 |
| - 26-35 | 40 | 22 | 1 Female 0 Male | 1/62 | 1.61 |
| - 36-45 | 27 | 14 | 1 Female 1 Male | 2/41 | 4.88 |
| - 46-60 | 23 | 14 | 1 Female 0 Male | 1/37 | 2.70 |
| - 60+ | 2 | 0 | 0 Female 0 Male | 0/2 | 0 |
| **Overall** | 139 | 68 | 207 | 7 | **3.38** |

The mean reaction scores (Table 4) for each assessment day during the induction phase remained low, indicating minimal irritation. Earliest reactions (Grade 1) were seen at assessment day 12. The lack of reactions until Day 12 indicates that initial exposures did not elicit any skin irritation.

**Table 4:** Mean reaction scores during induction phase. Day 22 (MU) refers to a designated make-up day for participants who missed an earlier assessment.

| **Assessment day** | **Mean reaction score (out of 5)** | **Standard deviation** |
| --- | --- | --- |
| Day 1 | 0.00 | 0.00 |
| Day 3 | 0.00 | 0.00 |
| Day 5 | 0.00 | 0.00 |
| Day 8 | 0.00 | 0.00 |
| Day 10 | 0.00 | 0.00 |
| Day 12 | 0.0048 | 0.07 |
| Day 15 | 0.024 | 0.16 |
| Day 17 | 0.029 | 0.17 |
| Day 19 | 0.034 | 0.18 |
| Day 22 (MU) | 0.045 | 0.21 |

### 3.4 Challenge Phase

There were no reactions during the Challenge Phase (Days 38 and 40) by any of the subjects to medical-grade *Picea abies* resin salve. Notably, participants who exhibited mild erythema (Grade 1) during the induction phase showed (Grade 0) no reactions during the challenge phase. No new cases of adhesive reactions were reported in this phase either. Sensitization would typically manifest as a reproducible reaction upon re-exposure during the challenge phase, which was not observed.

## 4 Discussion

The results of this study indicate that medical-grade *Picea abies* resin salve exhibits minimal irritation potential and does not induce sensitization in healthy adult volunteers. During the induction phase, only 3.38% of participants exhibited mild erythema (Grade 1) attributed to the resin salve. The results indicate that medical-grade *Picea abies* resin salve did not sensitize subjects nor lead to delayed contact allergy, based on the lack of reactions in the challenge phase. Although no sensitization reactions were observed, it cannot be ruled out (the rule of three, 3/n) (27) that up to 1.45 % of a larger population might be sensitized in a worst-case scenario. This also means that approximately 98.5% of the general population would not develop sensitization. These findings indicate that medical-grade *Picea abies* resin salve, is safe for topical application in terms of both irritation and sensitization potential. While allergic reactions may occur in highly predisposed patients (e.g., those with colophony allergy), the data supports that in the general population the risk is low.

Medical-grade *Picea abies* resin used in Abilar® is collected and purified without heat, reducing the formation of oxidized resin acids that are known to be more allergenic. Colophonium, on the other hand, contains high levels of abietic and dehydroabietic acids, which, while not sensitizers themselves, form skin-sensitizing oxidation products upon air exposure (15,16,23,24).

Colophonium used in technical products is often chemically modified, such as through reaction with maleic anhydride, producing maleopimaric acid (MPA), a potent contact allergen (28). Another modification, glycerol esterification, forms glyceryl 1-monoabietate (GMA), which also has sensitizing properties (29). Additionally, 13,14(beta)-epoxyabietic acid has been identified as a major allergen, with allergenicity similar to 15-HPA (28). Key allergens include 15-hydroperoxyabietic acid (15-HPA) and 7-oxo-dehydroabietic acid (7-O-DeA) (15,30–32). Low concentration of 7-oxo-dehydroabietic acid (33) is found also in medical-grade *Picea abies* resin.

The presence of this compound and other oxidized resin acids may explain the mild Grade 1 reactions observed in the induction phase, but further investigation is still required. However, the use of 10% medical-grade *Picea abies* resin in the salve, rather than higher concentrations, significantly reduces the overall amount of oxidized resin acids applied to the skin, further minimizing the risk of irritation or sensitization.

Sensitization to colophonium can develop progressively with repeated exposure. The oxidized resin acids can prime the immune system, eventually triggering an allergic reaction upon subsequent exposures (34). This process can lead to allergic contact dermatitis (ACD), which is clinically characterized by redness, itching, swelling, and, in more severe cases, blistering of the skin. Such reactions are commonly graded as Grade 2 or higher in HRIPT, with positive responses observed during the challenge phase confirming sensitization. The major contributing factor is the widespread usage of oxidized colophonium in various products, including adhesives, cosmetics, and topical treatments. Several studies of colophonium, propolis, and fragrance mixes suggest that contact with one of these allergens may increase the risk of developing delayed-type hypersensitivity reactions due to significant cross-reactivity with other chemically related compounds (35,36).

The findings of this study are particularly noteworthy when compared to the case series reported by Dendooven et al. (9), which highlighted instances of ACD in patients using Abilar® resin salve. In their study, six patients with leg ulcers developed ACD after applying the resin salve (9). The authors suggested that while oxidized resin acids are known culprits, other sensitizers present in the resin might also be responsible for allergic reactions. In the case series, the oxidation status of colophonium used in patch-testing of patients was not specified. Two patients did not react to colophonium but reacted to Abilar, while one patient reacted also to non-purified *Picea abies* resin in ethanol at 20%. The patients in Dendooven et al.’s study were polysensitized, reacting to multiple allergens including colophonium, abietic acid, *Myroxylon pereirae* (balsam of Peru), propolis, fragrance components, Compositae mix, limonene, linalool hydroperoxides, and Evernia furfuracea (9). All positive patch tests of these patients suggest a predisposition to hypersensitivity reactions, which may have contributed to their allergic responses to the *Picea abies* resin and Abilar®. In such individuals with an overactive or “primed” immune response, even trace amounts of otherwise low-risk compounds can elicit ACD.

In contrast, this study involved a larger cohort of healthy volunteers without known predispositions to multiple allergies or atopies, and these participants displayed minimal reactions. The findings align with Marzulli’s observations (37) that minimal irritation reactions are expected during induction, while true sensitization (an allergic response) manifests more frequently under prolonged exposure in predisposed populations. Moreover, the upper limit of exposure is specific to the HRIPT and does not reflect the limit for consumer exposure (38). This disparity reinforces the role of participant history and predisposition in sensitization outcomes, suggesting that medical-grade *Picea abies* resin salve is less likely to cause reactions in the among healthy adults’ population without pre-existing hypersensitivities under controlled use.

While HRIPT method used in this study effectively evaluates short-term irritation potential and sensitization, it has limitations for assessing long-term sensitization. As Kligman (39) noted, cumulative sensitization effects may not fully emerge within the test period. Longer-term studies may be needed to capture delayed sensitization responses. There are, however, clinical studies with longer usage of medical-grade *Picea abies* resin salve in clinical settings on wounds. In a prospective, randomized, controlled multicenter trial involving 37 patients (21 patients in medical-grade *Picea abies* resin group) with grade II-IV pressure ulcers. Complete healing of the pressure ulcers was significantly more common in the resin group (94% of the ulcers healed within 6 months). One of the patients experienced an allergic reaction, which was mild and resolved without medical intervention (8). In another study on leg ulcers in patients (19 patients in the medical-grade *Picea abies* resin salve group) with critical limb ischemia, the resin salve was compared with medical honey. Ulcers were fully healed clinically in 5 patients in the spruce resin group during the 6-month follow-up (83±47 days [range 30-183 days]). One patient in the medical-grade *Picea abies* resin salve showed local skin irritation at the ulcer site by the end of follow-up. An epicutaneous test ruled out colophony allergy, with a final diagnosis of infectious eczema confirmed by a consulting dermatologist (18). In another study, the healing rate of chronic, complicated surgical wounds with medical-grade *Picea abies* resin salve was 100% (23/23 patients). The mean ± SD healing time was 43 ± 24 days (range: 10–87 days; median: 41 days) (4). In an observational and clinical prospective follow-up study with 35 patients (19 in the medical-grade *Picea abies* resin salve group) that examined the healing of ordinary diabetic foot ulcers at homecare setting during the 145-day study period there were no cases of allergies or any signs of side effects (19). *Picea abies* resin acids have been used also for facial partial thickness burns of 19 children aged from 5 months to 6 years old. The treatment contributed to maintaining appropriate wound moisture, influencing the processes of granulation and epithelialization, and reducing pain during hospitalization. The average duration of *Picea abies* resin acids usage was 10.3±0.9 days. No adverse events were reported (40). Patients with acute wounds often require immediate and effective treatment with minimal risk of adverse reactions. The low incidence of reactions suggests that medical-grade *Picea abies* resin salve can be safely applied to acute wounds without significant concerns about irritation or sensitization. Chronic wounds require long-term management, and the risk of sensitization over prolonged use is a critical consideration. This study’s findings indicate that even with repeated exposure (as simulated in the induction phase), medical-grade *Picea abies* resin salve does not lead to significant sensitization in healthy volunteers. The absence of moderate to severe reactions (Grades 2-5) in the induction phase and no reactions in challenge phase suggests a low potential to cause allergic contact dermatitis.

In addition, in a guinea pig maximization test (GPMT) conducted using Freund’s Complete Adjuvant test (FCAT), no sensitization was observed (Test reference: CTL13/207-104TR, according to EN ISO 10993-10:2010). During the induction phase, intradermal injections with medical-grade *Picea abies* resin salve did not elicit any visible signs of sensitization. In the challenge phase, at a 100% concentration, both test and control groups showed 0/12 and 0/6 positive responses, respectively, at 24- and 48-hours post-application. The GMPT together with the absence of sensitization in the HRIPT, indicate that the medical-grade *Picea abies* resin is a safe for wound care in the general population.

As an ISO 13485–certified manufacturer, Repolar Pharmaceuticals Ltd. also maintains an ongoing post-market surveillance (PMS) system for Abilar®. This process includes collecting and evaluating reports from healthcare professionals, patients, and distributors regarding adverse events or product complaints. All cases are documented in accordance with regulatory guidelines, and periodic reviews ensure any new safety signals are rapidly addressed. Since Abilar® launch in 2008, 1.6 million tubes have been sold globally, primarily for various wound and burn treatments.

Given that most patients use a single tube per treatment course, this figure translates to well over 1.6 million individual patient applications. During this extensive real-world usage, 63 spontaneous adverse event reports were documented (as of January 2025). All of these reports involved mild to moderate, transient skin reactions, typically occurring in individuals with an underlying sensitivity to colophonium. Crucially, none of the reports have met the criteria for a serious adverse event, such as hospitalization or life-threatening complications. Based on sales data the overall reporting rate is approximately 0.004%, which is roughly 1 reported event per 25,000 tubes sold. This figure likely underestimates the true incidence of mild reactions, as many patients experiencing brief or self-resolving symptoms may not submit a formal report. Nonetheless, these PMS findings align with the minimal irritation and sensitization potential observed in this HRIPT study, further supporting the favorable safety profile of medical-grade *Picea abies* resin in real-world use.

## 5 Conclusion

This study demonstrates that medical-grade *Picea abies* resin salve (Abilar®) has minimal irritation potential and does not induce sensitization under controlled patch-test conditions in healthy volunteers. Only 3.38% of participants exhibited mild erythema (Grade 1) during the induction phase, and no reactions occurred in the challenge phase, indicating a low potential for allergic contact dermatitis in general population when used in accordance with its intended purpose. While the case series by Dendooven et al. highlight the occurrence of ACD in a small number of patients using Abilar®, this study provides evidence that such reactions are rare and generally mild in the broader population. Together with results from clinical trials, a guinea pig maximization test (GPMT), and extensive post-market surveillance showing rare instances of mild skin reactions (1 event per 25,000 tubes sold), these data support that Abilar® is safe for topical use in both acute and chronic wound settings for the general population. While rare allergic contact dermatitis cases have been reported in highly predisposed individuals, this study suggests that the product’s overall risk of causing irritation or sensitization is minimal. Routine clinical vigilance remains sensible, particularly for patients with known colophonium allergies.

## Data Availability

All data produced in the present study are available upon reasonable request to the authors

## Acknowledgments

The authors would like to thank all the volunteers who participated in this study. We also acknowledge the technical support provided by the staff at PCR Corp during the study. The study was sponsored by Repolar Pharmaceuticals Ltd.

## Author contributions

Kamilla Yamileva: Data curation, Formal Analysis, Writing – original draft, Visualization, and Writing – review & editing.

Evgen Multia: Conceptualization, Investigation, Methodology, Funding Acquisition, Project Administration, Data curation, Formal Analysis, Writing – original draft, Resources, Supervision, Visualization, and Writing – review & editing.

